# Effect of fasting therapy on vitamin D, vitality and quality of life. A randomized control trial

**DOI:** 10.1101/2022.04.08.22273614

**Authors:** Gulab Rai Tewani, Karishma Silwal, Gita Sharma, Dinesh Yadav, Aarfa Siddiqui, Sucheta Kriplani, Varsha Vijay Nathani, Neha Sharma, Jyoti Keswani, Himanshu Sharma, Pradeep M.K. Nair

**Affiliations:** Sant Hirdaram Yoga and Nature Cure Hospital, Bhopal; Yoga, Sant Hirdaram Medical College of Naturopathy & Yogic Sciences, Bhopal; Sant Hirdaram Medical College of Naturopathy & Yogic Sciences, Bhopal; Research, Sant Hirdaram Medical College of Naturopathy & Yogic Sciences, Bhopal

**Author notes:** Corresponding author **Corresponding authors Address:** Dr. Pradeep MK Nair, Professor & Head, Research, Sant Hirdaram Medical College of Naturopathy & Yogic Sciences, Sant Hirdaram Nagar, Bhopal-462020. Bachelor of Naturopathy and Yogic Sciences. Masters in Science. Doctorate in Philosophy. Masters in Public Health. **Author contribution statement:** Conceptualization: PMK, HS,GRT, KS, VVN, DY,; Data curation: PMK, SK, KS,KG, AS Formal analysis: PMK, KS,HS, GRT; Investigation: PMK,GS,MJ,SK,AS,DY,JK; Methodology: PMK, GRT, HS, KS,VVN,DY; Project administration: PMK, KS,KG, NS; Resources: GRT, HS; Supervision: PMK,HS, GRT; Visualization: PMK,GRT, HS; Writing - original draft: PMK; Writing - review & editing: PMK, KS, HS, GRT, SK,GS,JK,NS. **Funding:** No funds were received for this study. **Ethics approval** : The study was approved by the institutional ethics committee of Sant Hirdaram Medical College for Naturopathy & Yogic Sciences, Bhopal, India. **Consent to participate :** All the participants signed a written consent to express their consent to participate in the study. **Consent to Publish:** All the participants consented to publish their de-identified data on a medical journal. **Availability of data and material:** The data will be made available on request to the corresponding author.

**Keywords:** Vitamin D deficiency, Fasting, Diet, Therapeutic fasting, Vitality, Quality of life

## Abstract

**Background:** The aim of the present study was to determine the effects of prolonged fasting (10 days) in the vitamin D, B12 levels, body mass index (BMI), weight, hemoglobin, vitality and quality of life (QoL) compared to normal diet.

**Methods:** This randomized control trial included 52 participants (aged 19-74 years) randomized in to a fasting group (FG) or a normal diet group (NDG) with 26 participants in each group. The study was conducted at an in-patient setting where the FG were on a fasting diet (500 kCal/day) which included holy basil herbal tea, lemon honey juice and water (3 L). The NDG (1500 kCal/day) consumed routine diet that included Indian breads, pulses, steamed rice, vegetable salads and beverages.

**Results:** The FG has shown significant increase in the Vitamin D levels (p=0.003, d=0.475), vitality (p=0.006, d=0.425), physical QoL (p<0.001, d=0.549), psychological QoL (p=0.002, d=0.488), environmental QoL (p=0.004, d=0.457) compared to NDG. No significant changes were observed in Vitamin B12, weight, BMI, hemoglobin and social QoL. A weak to moderate (ρ= 0.330-0.483) positive correlation was observed between vitality scores and QoL domains, whereas BMI scores showed an inverse correlation (ρ=−0.280) with vitamin D levels.

**Discussion:** The results suggest that prolonged fasting can improve the vitamin D levels, vitality and promote quality life compared to normal diet. Unlike previous studies FG does not differ from NDG with respect to weight and BMI. Nevertheless, fasting may be utilized as an effective tool to tackle vitamin d deficiency and associated health insufficiencies.

**Trial Registry:** Clinical Trial Registry of India CTRI/2022/02/040446.

## Introduction

Therapeutic fasting is increasingly becoming popular among scientists, physicians and patients owing to its health benefits. The role of fasting in the prevention and management of various cardio-metabolic, musculoskeletal disorders are widely reckoned.(1)Majority of the reports on fasting have reported the efficacy of intermittent fasting (lasting for 16-48 hours) or calorie restriction but not on prolonged fasting (more than 4 days) barring few studies.(2,3) Prolonged fasting is defined as a medically supervised fasting regimen that lasts for a minimum of 7 days to 21 days with an average calorie intake of (200–500 kcal nutritional intake/day).(4) Fasting has potential therapeutic benefits as it has shown to reduce body weight,(5) body mass index (BMI), adiposity,(6) inflammation,(7) blood pressure, cholesterol, regulate blood sugar, insulin, glycated hemoglobin(8) etc. Hence fasting is viewed as a therapy with homoeostatic potential.

Vitamin D deficiency is reckoned as one of the growing public health issue worldwide reportedly affecting 30-80% of global population.(9)Similarly, the role of vitamin D from a mere micronutrient to an essential element in preserving health is been substantiated by numerous observational studies, which links vitamin D deficiency as one of prime precursor in cancers, immune disorders, metabolic disorders and neurodegenerative studies.(10)While Vitamin D supplementation is considered as a probable measure in alleviating Vitamin D deficiency, previous data suggests that a hypocaloric diet induced weight loss can improve the vitamin D status among patients.(11)Recently Arankale et al and Żychowska et al has reported that fasting can improve the circulating levels of vitamin D and has an impact on its metabolites as well.(3,12)However, both these studies were conducted on healthy volunteers without any controls.

It will be interesting to study the impact of prolonged fasting on the vitamin D levels of participants with different diseases, as fasting is widely practiced both as a therapeutic, cultural and religious practice. Further, the association of prolonged fasting with vitality and quality of life is an unexplored domain. Fewer studies have reported fasting to act as a mood enhancer in pain,(8) moderately relieve stress, anxiety, depression(13) and offers rewarding psychological experiences.(14) However, all the previous reports necessitate the need for future randomized controlled trials. Further barring Ramadan fasting and calorie restriction, the impact of prolonged fasting on health indices like weight, BMI etc were not studied on individuals with various medical co-morbidites.

The study hypothesized that a 10 days prolonged fasting program will increase the vitamin D levels, Vitamin B12 levels, enhance the vitality, quality of life, reduce body weight and BMI.

## Methods

### Study settings and design

This study was conducted at a private yoga and naturopathy medical college hospital in India. The study approved by institutional ethics committee via (F.No:12/SHMCNYS-IEC/P45/2021-2022) and was registered in Clinical Trial Registry of India CTRI/2022/02/040446. Written informed consent was obtained from all participants prior to participation in the study. This was a pragmatic, parallel group, randomized controlled trial (n=52). Eligible participants were randomized by the study coordinator at a ratio of 1:1 using computer random software in to fasting arm or normal diet arm. Figure 1 depicts the trial profile.

**Figure 1:**
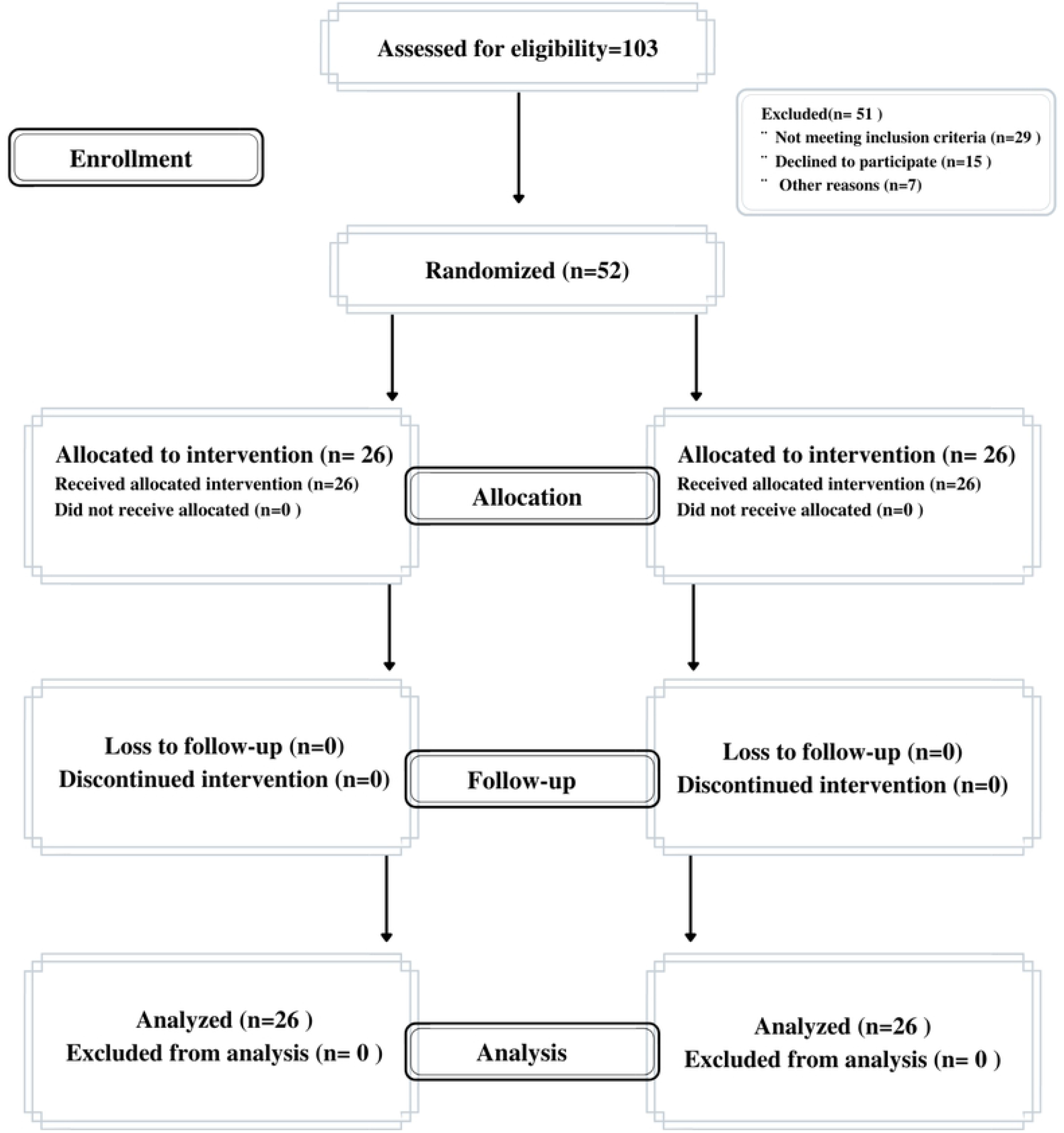
Trial Profile.

### Study Participants

The study participants were those volunteers with diverse medical conditions enrolled for a 10 days lifestyle modification program at the in-patient department of a private yoga and naturopathy medical college hospital. Both men and women aged between 20 to 75 years participated in this program. All the patients who were underweight, with known nutritional deficiency, psychiatric disorders were excluded from participating in this study. Any participants who have a previous history of fasting for more than 10 days in the last 6 months were also excluded from the study. Participants were also excluded if they are a part of any other clinical trial or following any dietary regimens/food supplements.

### Sample size

Based on a previous pilot study,(3) for an effect size of 0.8, level of significance (α)=0.05 and power 80% the sample size was calculated to be n=40 by using G Power software. Considering a drop out of 30% the final sample size was n=52 (n=26 in each arm).

### Interventions

The fasting arm received 10 days of fasting regimen which includes two days of feeding (preparation day 1 and re-feeding day 10) that included herbal tea (100 mL), lemon honey juice(100 mL), cucumber juice (200 mL), boiled green grams (75gms), Rice and lentil porridge (100 g), Plain vegetable soup (150 mL), Water (3 L) at different intervals during the day, which amounts for 1000 kCal/day approximately. For the rest of 8 days (day 2 to day 9) the participants were on a fasting diet (500 kCal/day) which included holy basil herbal tea (100 mL), lemon honey water (1,000 mL), and water (3 L). The normal diet arm consumed routine diet that included Indian breads, pulses, steamed rice, vegetable salads and beverages that amounts to 1500 kCal/day. In addition to the dietary regimen both the study group and the control group received yoga and naturopathy therapies like hip bath, cold water enema, and mud packs to the abdomen and eyes which are intended to promote elimination.(3)

### Assessments

Blood samples were collected from both the study group at baseline and at the 10^th^ day after intervention to evaluate the changes in the serum levels of vitamin D, vitamin B12 and hemoglobin. Additionally, we evaluated the changes in weight, body mass index, quality of life and vitality. World Health Organization Quality of Life (WHO-QoL BREF)(15) instrument was used to assess the quality of life, whereas vitality (defined as the innate power to heal and to be resilient)(16) was measured on a scale of 1 to 10, using a visual analogue scale where ‘0’ represents poor vitality and ‘10’ represents higher vitality.

### Statistical analysis

All the data were analyzed using Jeffreys’s Amazing Statistics Program (JASP) version 0.16. Shapiro’s Wilk’s test was used to test the normality of the data. Wilcoxon sign-rank test (within the group testing) and Mann-Whitney test (between the group testing) were used to analyze the non-normally distributed data, where as paired T test and Independent T test were used for analyzing the normally distributed samples. A P-value of 0.05 is considered as a significant change.

## Results

Of the 103 individuals screened for the trial, n=52 were randomized to the fasting (n=26) or control groups (n=26). All the 52 participants completed the study without any adverse events. The baseline characteristics are tabulated in table 1. The changes in the body weight (p= 0.51), BMI (P=0.99), vitamin B12 (P=0.40) and hemoglobin (P=0.77) were not significant in the fasting group compared to the control group. However, a within the group analysis has shown that the changes in weight and BMI were significant (P < .001) before and after the intervention in both the fasting and control group. However, the changes in vitamin B12 and hemoglobin remained non-significant in both the groups. The mean changes in these parameters from the baseline to end point are tabulated in table 2.

**Table 1:** Baseline demographic characteristics of the participants.

| **Parameters assessed** | **Fasting Group (n=26)** | **Control group(n=26)** |
| --- | --- | --- |
| **Age** | 53.57±10.21 | 49.96±15.62 |
| **Sex** | Males-11<br>Females-15 | Males-11<br>Females-15 |
| **Height** | 159.65±9.68 | 158.92±9.55 |
| **Weight** | 67.9±10.53 | 70.88±28.46 |
| **Body Mass Index** | 26.86±4.71 | 27.8±9.11 |
| **Socioeconomic status** | Upper Class-9<br>Middle class-17 | Upper class-1<br>Upper Middle class-2<br>Middle class- 23 |
| **Habits** | Smoking-1 | Smoking-1 |
| **Medications** | Under Medications-9<br>No Medication-17 | Under Medications-14<br>No Medication-12 |
| **Conditions** | Diabetes-2<br>Dyslipidemia-1<br>Gastrointestinal disorders-2<br>Hypertension-5<br>Hyperthyroidism-1<br>Hypothyroidism-1<br>Insomnia-1<br>Lumbago-3<br>Osteoarthritis-2<br>Obesity-5<br>Skin disorders-3 | Asthma-2<br>Diabetes-6<br>Fatty Liver-1<br>Gastrointestinal disorder-3<br>Hypertension-5<br>Lumbago-1<br>Migraine-1<br>Osteoarthritis-3<br>Obesity-4 |

**Table 2:** Comparison of variables between the fasting and control group.

| Variables assessed | Fasting group |  | Within group analysis p value | Normal diet group |  | Within group analysis p value | Between group analysis p value | Effect size |
| --- | --- | --- | --- | --- | --- | --- | --- | --- |
|  | Pre (Mean±SD) | Post (Mean±SD) |  | Pre (Mean±SD) | Post (Mean±SD) |  |  |  |
| **Weight (kg)** | 67.9±10.53 | 65.38±10.64 | <0.001 | 70.88±28.46 | 68.68±28.16 | <0.001 | 0.57^ | 0.155 |
| **Body mass index (kg/m<sup>2</sup>)** | 26.86±4.71 | 25.99±4.53 | <0.001 | 27.8±9.11 | 26.92±8.99 | <0.001 | 0.63^ | 0.131 |
| **Vitamin D (ng/ml)** | 25.69±10.66 | 31.5±11.13 | <0.001 | 20.92±8.89 | 22.4±11.29 | 0.23 | 0.003^ | 0.475 |
| **Vitamin B12 (pg/ml)** | 384.42±336.43 | 365.5±207.92 | 0.75 | 355.91±224.9 | 358.26±140.51 | 0.25 | 0.40^ | 0.136 |
| **Hemoglobin (gm/dl)** | 13.12±1.14 | 13.04±1.09 | 0.91 | 12.9±1.39 | 12.95±1.34 | 0.72 | 0.77* | 0.07 |
| **Vitality** | 6.73±1.18 | 7.61±1.02 | <0.001 | 5.42±1.39 | 8.46±0.9 | <0.001 | 0.006^ | 0.425 |
| **WHO-Quality of life (QoL)** |  |  |  |  |  |  |  |  |
| **Physical QoL** | 70.38±14.45 | 82.5±11.85 | <0.001 | 55.61±17.23 | 68.8±13.64 | <0.001 | <0.001^ | 0.549 |
| **Psychological QoL** | 71.03±15.01 | 83.26±14.09 | <0.001 | 59.46±15.17 | 70.73±15.66 | 0.001 | 0.002^ | 0.488 |
| **Social QoL** | 78.46±13.6 | 82.88±13.56 | 0.02 | 72.15±18.7 | 73.34±19.53 | 0.42 | 0.06^ | 0.302 |
| **Environment QoL** | 84.53±11.34 | 88.26±11.88 | 0.01 | 71.53±16.05 | 77.92±13.69 | 0.006 | 0.004^ | 0.457 |

\*Students T test; ^Mann-Whitney U test, QoL-Quality of Life

Significant increase in the levels of vitamin D (P=0.003) was observed in fasting group compared to the control group. Similarly, statistically significant increase was observed in vitality scores (P= 0.003) of the fasting group when compared to the controls. A within group analysis between the mean values before and after the intervention has shown a significant rise in the vitamin D levels (P < .001) and vitality (P < .001) in the fasting group. However, the control group has shown significant increase only in the vitality scores (P < .001) whereas the changes in vitamin D in this group remained insignificant (P=0.23). The changes are depicted in figure 2.

**Figure 2:**
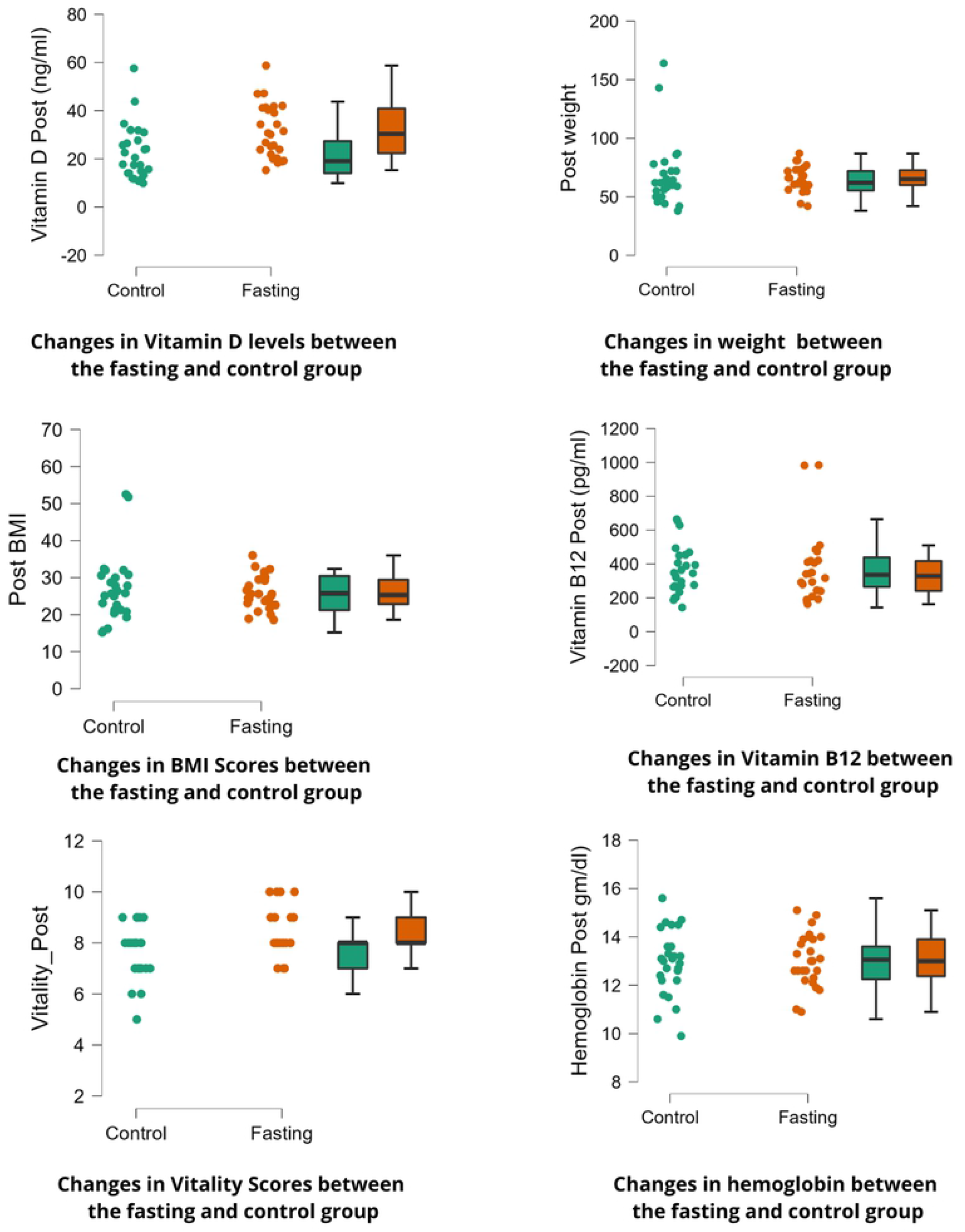
Changes in the mean values of outcome measures between the fasting and control group.

All the participants in the fasting group has shown significant improvement in the quality of life domains of WHOQOL namely physical domain (P < .001), psychological domain (P=0.002), environmental domain (P=0.004) except social domain (P=0.06) compared to their controls. Nevertheless, a within group analysis has shown significant improvement in all the QOL domains in the fasting group and control group, except for psychological domain (P=0.09) in the control group. The detailed results are tabulated in table 2.

Spearman’s correlation has shown a weak negative correlation between BMI and Vitamin D levels (ρ=−0.280,P=0.04). Vitality status was reported to have a moderate positive correlation with physical QoL(ρ= 0.442,P=0.001) and environmental QoL (ρ= 0.483,P< .001). Further vitality status also demonstrated to have a weak positive correlation with psychological QoL (ρ= 0.348, P=0.001) and social QoL ((ρ= 0.330, P=0.01). The correlations between these variables are plotted in figure 3 and 4.

**Figure3:**
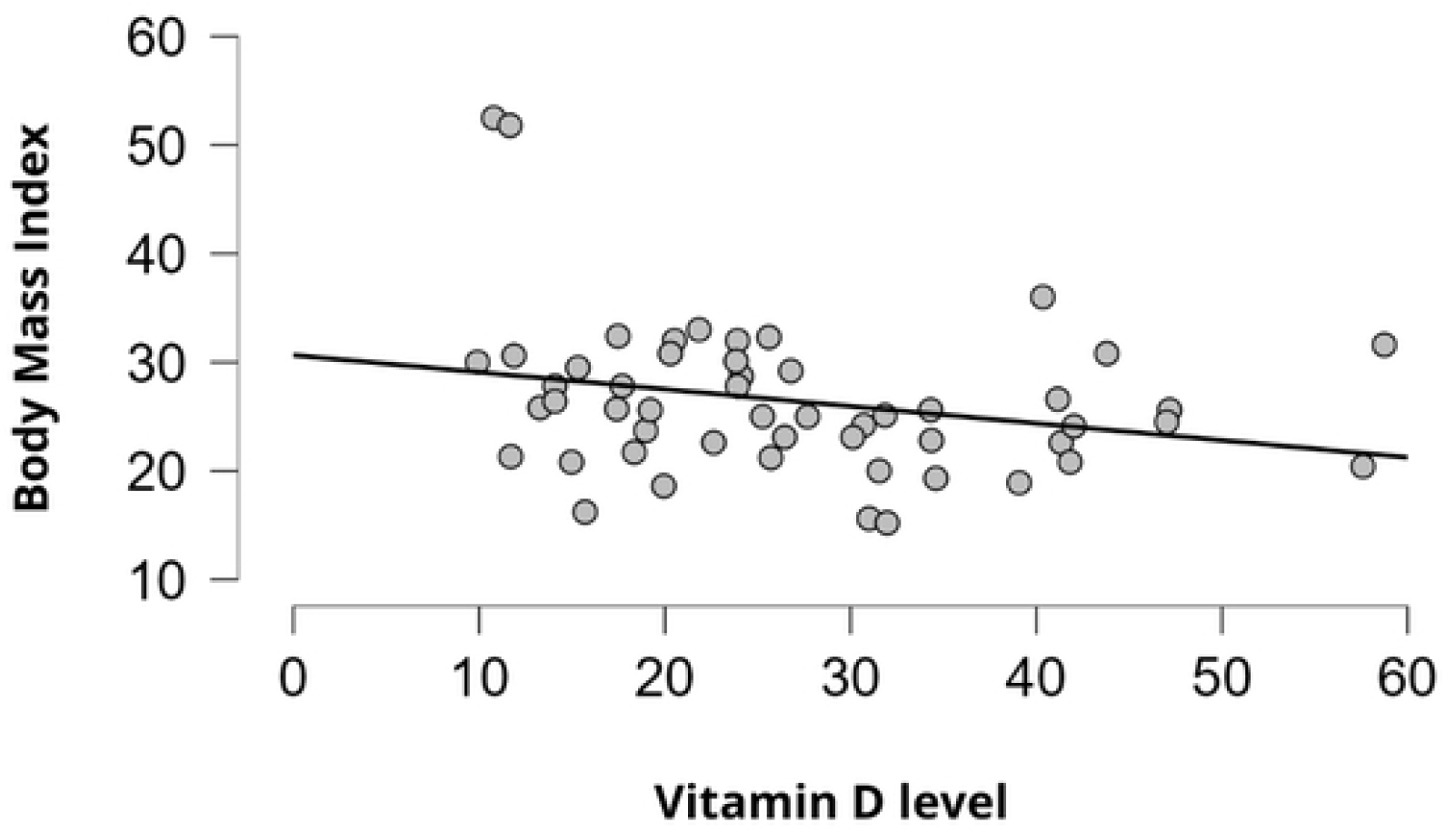
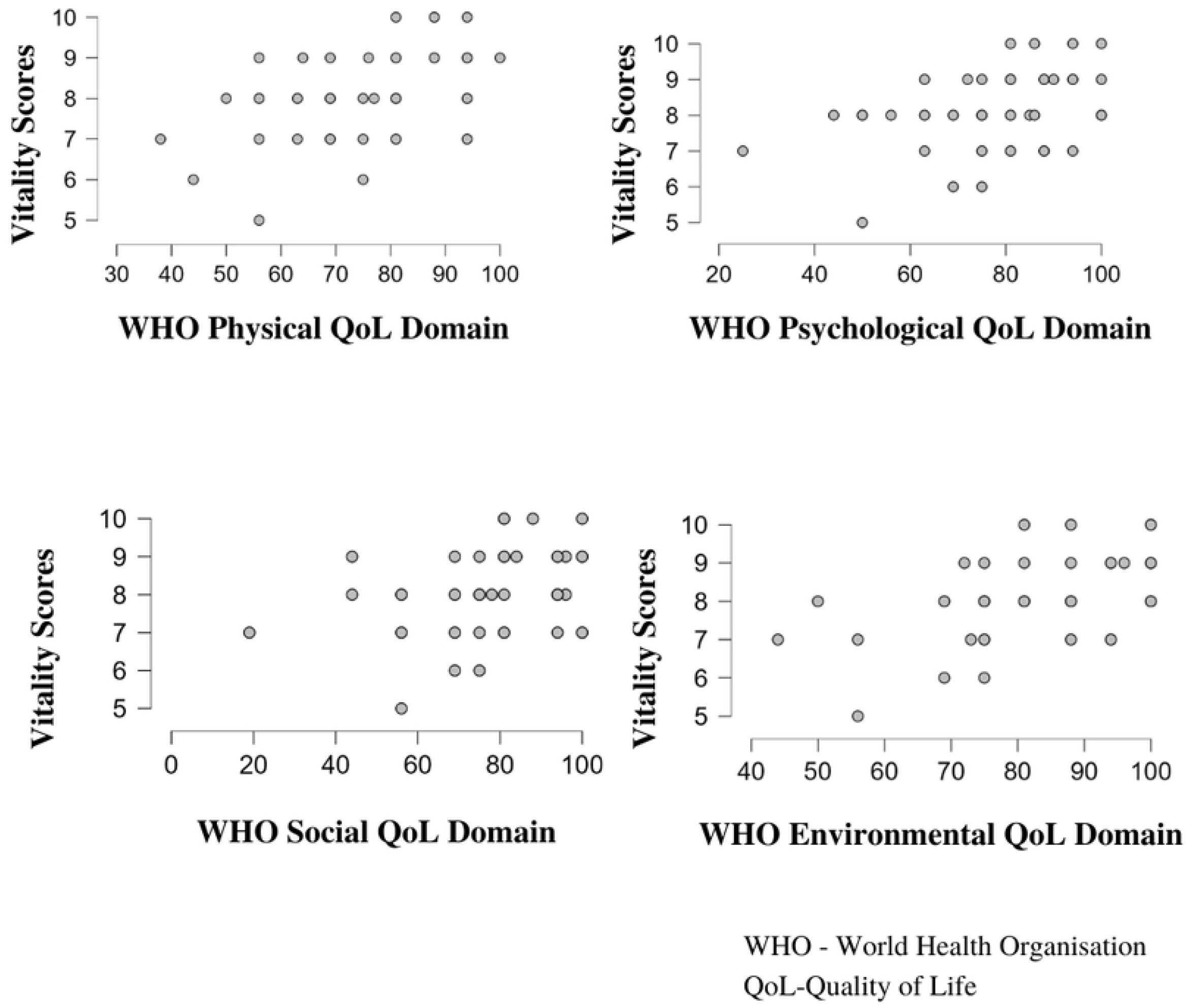
Relationship between Vitamin D level and Body mass index.

**Figure 4:** Correlation between Vitality and Quality of life.

## Discussion

This study was conducted to validate the findings of our previous pilot study which reported 10 days fasting significantly improve the levels of vitamin D and B12, weight reduction, reduces BMI and prime the other physiological functions among female healthy volunteers.(3) This randomized controlled trial indicates a positive role of fasting therapy over the vitamin D levels, quality of life and vitality of the patients. The present study demonstrated an increase in the vitamin D levels after fasting compared to normal diet. This is strengthening the observations from earlier studies which reported 8-10 days of fasting can increase the levels of serum vitamin D.(3,12)

Earlier studies postulate that a reduction in body weight during fasting and/or exercise as the rate limiting factor that stimulates the redistribution of vitamin D stored in the adipose tissue and skeletal muscles.(3,11,12,17) In the present study, participants from both the groups had reported to have significant reduction in their weight and BMI. Compared to the baseline, fasting group had an average of 3.7% reduction in weight and 0.25% reduction in the BMI scores, whereas the control group had 3.1% and 0.23% reduction in weight and BMI respectively.

Statistically fasting group did not differ from that of the control group with respect to weight and BMI scores. This is in contrary to the results reported by a recent systematic review that has shown intermittent fasting to reduce body weight.(18) A previous observational study reported significant reduction in weight on a 20 day fasting program.(2) However, the absence of a control group in this study limits its generalizability. To our knowledge this is the first randomized control trial to compare the impact of prolonged fasting (10 days) with normal diet on body weight in a controlled residential setting.

The present data suggests that the changes in vitamin D level post fasting may be independent of reduction in weight or BMI. However, a correlation analysis suggests that reduction in BMI was associated with an increase in the vitamin D levels. This is strengthening the earlier reports that vitamin D levels are inversely proportional to the body weight and BMI.(19) Adequate vitamin D levels are warranted in humans, as vitamin D is necessitated in all major functions of the body and mind.(20) Reduced levels of vitamin D predisposes to obesity, insulin resistance and other metabolic disorders.(21) Further optimum levels of vitamin D is considered as a pre-requisite in treating and preventing diseases ranging from diabetes to cancer.(22)As on date vitamin D deficiency and insufficiency is a global concern. In this scenario fasting therapy may be a promising solution for not only mitigating vitamin D deficiency but also can prevent its ramifications.

We observed 7% reduction in the vitamin B12 levels in the fasting group, whereas the control group had an increase of 1% in their vitamin B12 levels. However, these changes were not significant. The present study contradict the results of the previous pilot study which reported fasting to increase the vitamin B12 levels on a group of healthy volunteers.(3)Vitamin B12 shares an inverse relationship with weight,(23) the insignificant changes in the weight of the study participants may be a reason for the insignificant changes in vitamin B12 levels. Nevertheless, this needs to be revalidated by future studies.

Another significant change observed in the present study was the improvement in the vitality status and the quality of life in the fasting group. Vitality is reckoned as one of the important component in measuring one’s abilities irrespective of his/her disabilities.(24)There is a growing interests in measuring the vitality as one of the indicator of well-being among various medical disciplines.(25–27)Similarly, improving the quality of life remains as one the primarily sought endpoint in health care delivery and research.(28)

The present data suggests a compelling positive relationship between the vitality status and the quality of life. Therefore, understanding and prodding the vitality status among the patients may be a key factor to be considered in future research and clinical practice, as the paradigm of health care and research is shifting towards a patient-centric approach. However, there is a need for a better scale to measure vitality as VAS scoring may not return an accurate measure.

### Limitations of the study

The status of vitamins D, B12, weight, BMI, vitality and quality of life were not measured post fasting program. This remains as one of the limitations of this study as the changes in these parameters during re-feeding phase in an uncontrolled atmosphere will add more information about the sustainability of the results. Further, due to operational constraints at the clinical setting, the authors could not control for the difference in disease status and medication use of the participants, which may be viewed as a potential confounder in this study. The authors have not assessed the specific disease related effects of fasting in these participants which may be included in future studies. Irrespective of the limitations, this is the first randomized controlled trial to report an association between therapeutic fasting and vitamin D.

## Conclusion

Prolonged fasting up to 10 days has demonstrated to be safe and helps in improving the vitamin D levels, vitality and quality of life of patients. This may have considerable preventive and prophylactic benefits. Future studies should evaluate the long-term effects and cost effectiveness of fasting as a public health tool.

## Data Availability

All relevant data are within the manuscript and its Supporting Information files.

